# Age and product dependent vaccine effectiveness against SARS-CoV-2 infection and hospitalisation among adults in Norway: a national cohort study, January – September 2021

**DOI:** 10.1101/2021.11.12.21266222

**Authors:** Jostein Starrfelt, Eirik Alnes Buanes, Lene Kristine Juvet, Trude Marie Lyngstad, Gunnar Øyvind Isaksson Rø, Lamprini Veneti, Hinta Meijerink

**Affiliations:** Department of Infection Control and Preparedness, Norwegian Institute of Public Health, Oslo, Norway; Norwegian Intensive Care and Pandemic Registry (NIPaR), Helse Bergen Health Trust, Bergen, Norway; Department of Anaesthesiology and Intensive Care Haukeland University Hospital, Bergen, Norway; Department of Infection Control and Vaccines, Norwegian Institute of Public Health, Oslo, Norway; Department of Development and Analytics, Norwegian Institute of Public Health, Oslo, Norway

## Abstract

**Background:** SARS-CoV-2 vaccines show high effectiveness against infection and (severe) disease. However, few studies estimate population level vaccine effectiveness against multiple COVID-19 outcomes, by age and including homologous and heterologous vaccine regimens.

**Methods:** Using Cox proportional hazard models on data from 4 293 544 individuals (99% of Norwegian adults), we estimated overall, age-, and product-specific vaccine effectiveness against SARS-CoV-2 infection, hospitalisation, ICU admission and death in Norway, using data from national registries. Vaccine status was included as time-dependent variable and we adjusted for sex, pre-existing medical conditions, country of birth, county of residence, and crowded living conditions.

**Findings:** Adjusted vaccine effectiveness among fully vaccinated is 72·1% (71·2–73·0) against SARS-CoV-2 infection, 92·9% (91·2–94·2) against hospitalisation, 95·5% (92·6–97·2) against ICU admission, and 88·0% (82·5–91·8) against death. Among partially vaccinated, the effectiveness is 24·3% (22·3–26·2) against infection and 82·7% (77·7–86·6) against hospitalisation. Vaccine effectiveness against infection is 84·7% (83·1-86·1) for heterologous mRNA vaccine regimens, 78·3% (76·8-79·7) for Spikevax (Moderna; mRNA-1273), 69·7% (68·6-70·8) for Comirnaty (Pfizer/BioNTech; BNT162b2), and 60·7% (57·5-63·6) for Vaxzevria (AstraZeneca; ChAdOx nCoV-19; AZD1222) with a mRNA dose among fully vaccinated.

**Interpretation:** We demonstrate good protection against SARS-CoV-2 infection and severe disease in fully vaccinated, including heterologous vaccine regimens, which could facilitate rapid immunization. Partially vaccinated were less likely to get severe disease than unvaccinated, though protection against infection was not as high, which could be essential in making vaccine prioritisation policies especially when availability is limited.

**Funding:** Norwegian Institute of Public Health, Helse Bergen Health Trust

## Background

In December 2019, cases of acute respiratory syndrome were reported in Wuhan City, Hubei province, China, and quickly was discovered to be caused by a novel coronavirus, severe acute respiratory syndrome coronavirus 2 (SARS-CoV-2). Coronavirus disease 2019 (COVID-19) spread rapidly and was declared a pandemic on 11^th^ of March 2020 by the World Health Organization. The pandemic challenged health care systems and has only partially been abated by non-pharmaceutical control measures. Therefore, effective vaccines would be essential to control the outbreak, and many vaccines were rapidly developed. The first vaccines were approved for large scale use in December 2020, including Comirnaty (Pfizer/BioNTech; BNT162b2), Spikevax (Moderna; mRNA-1273), Vaxzevria (AstraZeneca; ChAdOx nCoV-19; AZD1222), and Janssen (Johnson & Johnson; Ad26.COV2.S). Traditionally, a single vaccine is administered in multiple doses as homologous regimen. However, many countries adopted more flexible policies allowing “mixing and matching” of vaccines during SARS-CoV-2 vaccination campaigns, usually in response to supply constraints or policy changes, this strategy is termed “heterologous” regimen.^1,2^

COVID-19 vaccines have exhibited good protection against SARS-CoV-2 disease, and the vaccine effectiveness increases with the severity of the outcome - effectiveness against infection is lower than hospitalization, intensive care unit admission (ICU) and death.^3-7^ A recent meta-analysis estimated an overall effectiveness of 83% against SARS-CoV-2 infection after two doses, with higher protection from mRNA vaccines.^8^ However, effectiveness may differ among virus variants and product types, as well as being impacted by dose intervals or population structure (age distribution, risk groups).^9-15^ In addition, waning of vaccine-induced immunity is a concern that could result in lower vaccine effectiveness as time passes. ^16,17^ Determining the effectiveness of vaccines on population level after roll-out of COVID-19 vaccination programmes as well as comparison between vaccine types is essential for vaccine implementation and guidelines.

By 27^th^ of September 2021, a total of 187 282 SARS-CoV-2 cases were reported to the Norwegian Surveillance System for Communicable Diseases (MSIS) in a population of approximately 5.3 million, with 5 193 individuals hospitalised due to COVID-19 and 861 COVID-19 associated deaths. From early February 2021, the Alpha variant (B1.1.7) was the dominant circulating strain in Norway, being replaced by the Delta variant (B.1.617.2) by July 2021.^11,14,15^ On 27^th^ of December 2020, Norway started COVID-19 vaccination, initially targeted towards elderly (>65 years) and risk groups. Of those ≥18 years, 84% were considered fully vaccinated by 27^th^ of September, with the majority receiving a mRNA vaccine regimen; Comirnaty, Spikevax, or a heterologous combination of both (accepted since June 2021).^18^ Vaxzevria was included in the Norwegian national vaccine programme until 11^th^ of March 2021; those who received one dose were offered a second dose with a mRNA vaccine.

The purpose of this study is twofold – to quantify the general vaccine effectiveness achieved in the Norwegian population, with the realized mixture of vaccine types, current population characteristics and epidemic trajectory and to directly estimate vaccine effectiveness across vaccine types and age-groups. To understand the impact of COVID-19 vaccination in the Norwegian population, we aim to estimate the vaccine effectiveness against SARS-CoV-2 infection, hospitalisation with COVID-19 as main cause, intensive care unit (ICU) admission and death with COVID-19, specifically looking at differences between age and vaccine regimen. We hypothesize that vaccine effectiveness declines with age and varies among vaccine regimens.

## Methods

### Study population

For this population-based cohort study, we obtained data from the Norwegian National Preparedness registry for COVID-19 (BeredtC19) (Supplementary table S1), which contains individual-level data from national central health registries, national clinical registries, and other national administrative registries. This enabled us to follow-up the whole adult population over time as well as correct for various factors otherwise only available in specific research studies or only on an aggregated level. We included all adult Norwegian individuals (≥18 years by end 2021) with a valid national identity number and registered in the National Population Registry as living in Norway.

We excluded individuals for which the interval between first and second dose was shorter than the minimum intervals (1 546 individuals), all individuals with more than two doses registered before the 27^th^ of September 2021 (15 966 individuals) as well as those with unknown vaccine types (346) or vaccine type not part of the Norwegian vaccination program (Janssen – 4 381, Covishield – 19). The minimum interval between doses, based on the vaccine type used for the first dose, was 19 days for Comirnaty, 22 days for Spikevax, and 21 days for Vaxzevria. In addition, we excluded individuals registered with an infection prior to 1^st^ of January 2021 (40 118). Individuals were fully excluded from the dataset, also while unvaccinated. Data were extracted from the registries on the 4^th^ of November 2021. Number of infections and hospitalisations as well as distribution of vaccination status over time are available in Supplementary figures S1 and S2).

### Definitions

We defined SARS-CoV-2 infection as a positive SARS-CoV-2 PCR test reported to MSIS. We used the Norwegian Intensive Care and Pandemic Registry (NIPaR) to identify individuals who were hospitalised or admitted to an ICU. We included all hospital admissions where COVID-19 was registered as the main cause for admission regardless of length of stay. We defined ICU admission as individuals who tested positive for SARS-CoV-2 and were admitted to an intensive care unit (≥24 hours), required mechanical ventilatory support (invasive or non-invasive), or persistent administration of vasoactive medication. All COVID-19 associated deaths are registered in MSIS and defined as anyone with a positive SARS-CoV-2 PCR test who died with COVID-19 as main or underlying causes in the Cause of Death Register (DÅR) or those notified directly to MSIS as a COVID-19 associated death. We use testing date as time of infection (positive PCR test) for all outcomes to determine vaccine status.

Individual vaccination histories were generated from the Norwegian Immunisation Registry (SYSVAK), including date of vaccination and type of vaccine. Vaccine type was Comirnaty, Spikevax, heterologous mRNA regimen, Vaxzevria, or Vaxzevria in combination with an mRNA vaccine.

Vaccination status was defined as:

- Unvaccinated: unvaccinated up to 21 days after first vaccine dose
- Partially vaccinated: ≥21 days after first vaccine dose up to seven days after second vaccine dose
- Fully vaccinated: seven days or more after second vaccine dose

To adjust for confounding, several covariates were included in our analysis. County of residence based on the National Population Registry, coded as living in the capital or surrounding region (Oslo or Viken) versus the rest of Norway, was included as this region experienced higher infection rates and was prioritised for vaccination from March to July 2021. We included country of birth as recorded in National Population Registry (Norway, abroad or unknown) and crowded living conditions as registered in Statistics Norway (crowded, not crowded, or unknown) since both are potentially associated with vaccine coverage and risk of infection. Individuals with pre-existing medical conditions associated with moderate or high risk of serious COVID-19 illness were prioritised for vaccination and was also included in the adjusted model. Data sources, variables, and detailed description of crowding and risk groups can be found in the supplementary information. Missing values were considered as a separate category for each of the variables. Ethical approval was granted by Regional Committees for Medical and Health Research Ethics (REC) South East (reference number 122745).

### Data analyses

We estimated the vaccine effectiveness with Cox proportional hazards models, using vaccine status as time-varying covariate for all individuals included. We right-censored individuals at the time of an event (SARS-COV-2 infection, hospitalisation, ICU admission or death associated with COVID-19), time of death (all cause) or end of follow-up period (27^th^ of September). Vaccine effectiveness is defined as 100*(1 –β), with β the proportional hazard associated with vaccine status.

For crude vaccine effectiveness estimates, we only used vaccine status as time-varying covariate with no adjustments for covariates. For adjusted vaccine estimates, we implemented stratified analyses using *strata(variable)* in the *survival-*package. The adjustments made using stratification in the survival package assumes that the impact of the adjustment variables is non-proportional. In essence for a given factor, all levels are assumed to have their own baseline hazard-rate over time. Vaccine status is implemented as a regular parameter and assumes proportionality. We adjust using stratification instead of fixed covariates since the infection risk is highly dependent on these covariates through time, though not in the same manner over the whole study period, and that the Norwegian vaccine programme has prioritised these groups differentially and/or there are reported differences in the degree to which vaccines are accepted within the categories. This adjustment is the most generic and imposes fewer restrictive assumptions on the impact of the covariates.

To estimate specific vaccine effectiveness for age-groups we implemented independent Cox-models still adjusting for the remaining covariates as *strata*. Vaccine status was implemented either by combining all vaccine types (thus assuming similar effectiveness across vaccines) to estimate a population level vaccine effectiveness of the vaccination program in Norway or by implementing vaccine status as the combination of vaccine-type and vaccine status. All models were implemented in the statistical software *R*^19^, using the survival package.^20^

## Results

In total, 4 293 544 individuals were included, of whom 3 843 420 (90 %) received at least one vaccine dose during the study period. Of those, 88 618 (2.3 %) were diagnosed with SARS-CoV-2 infection, 2 752 (0.07 %) were hospitalised with COVID-19 as main cause for admission, 530 (0.01 %) were admitted to the ICU, and 323 (0.008 %) died with COVID-19. Overall, the median follow-up time was 269 days (inter quartile range (IQR): 269-269), for those with SARS-CoV-2 infection it was 134 days (IQR: 71-220), and 148 days (IQR: 73-207) for individuals censored by death not associated with COVID-19. Characteristics of the study population and by outcome can be found in Table 1.

**Table 1.** Characteristics of study population and by outcomes of interests; SARS-CoV-2 infections, hospitalisation, ICU admission and death, Norway, 1^st^ January 2021 – 27^th^ September 2021

|  |  | Total |  | SARS-CoV-2 infection |  | Hospitalisation |  | ICU admission |  | COVID-19 deaths |  |
| --- | --- | --- | --- | --- | --- | --- | --- | --- | --- | --- | --- |
|  |  | n | % | n | % | n | % | n | % | n | % |
| **All** | - | 4 293 544 | 100·0 | 88 618 | 100·0 | 2 752 | 100·0 | 530 | 100·0 | 322 | 100·0 |
| **Age groups** | 18-24 years | 452 535 | 10·5 | 21 675 | 24·5 | 59 | 2·1 | 4 | 0·8 | 0 | 0·0 |
|  | 25-34 years | 739 120 | 17·2 | 20 612 | 23·3 | 231 | 8·4 | 25 | 4·7 | 1 | 0·3 |
|  | 35-44 years | 701 643 | 16·3 | 17 924 | 20·2 | 416 | 15·1 | 46 | 8·7 | 4 | 1·2 |
|  | 45-54 years | 732 365 | 17·1 | 15 561 | 17·6 | 700 | 25·4 | 131 | 24·7 | 13 | 4·0 |
|  | 55-64 years | 651 977 | 15·2 | 7 737 | 8·7 | 571 | 20·7 | 133 | 25·1 | 33 | 10·2 |
|  | 65-74 years | 541 512 | 12·6 | 3 041 | 3·4 | 416 | 15·1 | 112 | 21·1 | 59 | 18·3 |
|  | 75-84 years | 339 022 | 7·9 | 1 385 | 1·6 | 274 | 10·0 | 72 | 13·6 | 95 | 29·5 |
|  | ≥85 years | 135 370 | 3·2 | 683 | 0·8 | 85 | 3·1 | 7 | 1·3 | 117 | 36·3 |
| **Sex** | Male | 2 156 029 | 50·2 | 46 567 | 52·5 | 1 654 | 60·1 | 357 | 67·4 | 186 | 57·8 |
|  | Female | 2 137 515 | 49·8 | 42 051 | 47·5 | 1 098 | 39·9 | 173 | 32·6 | 136 | 42·2 |
| **Underlying conditions** | No risk group | 3 381 800 | 78·8 | 77 234 | 87·2 | 1 629 | 59·2 | 257 | 48·5 | 96 | 29·8 |
|  | Medium risk | 797 225 | 18·6 | 10 276 | 11·6 | 914 | 33·2 | 218 | 41·1 | 149 | 46·3 |
|  | High Risk | 114 519 | 2·7 | 1 108 | 1·3 | 209 | 7·6 | 55 | 10·4 | 77 | 23·9 |
| **County of residence** | Capital + surrounding region (Oslo & Viken) | 2 748 290 | 64·0 | 36 131 | 40·8 | 1 001 | 36·4 | 189 | 35·7 | 107 | 33·2 |
|  | Other | 1 545 254 | 36·0 | 52 487 | 59·2 | 1 751 | 63·6 | 341 | 64·3 | 215 | 66·8 |
| **Country of birth** | Norway | 3 194 256 | 74·4 | 52 202 | 58·9 | 1 214 | 44·1 | 244 | 46·0 | 200 | 62·1 |
|  | Outside Norway | 782 914 | 18·2 | 34 775 | 39·2 | 1 379 | 50·1 | 255 | 48·1 | 69 | 21·4 |
|  | Unknown | 316 374 | 7·4 | 1 641 | 1·9 | 159 | 5·8 | 31 | 5·8 | 53 | 16·5 |
| **Crowding** | Yes | 333 448 | 7·8 | 17 491 | 19·7 | 488 | 17·7 | 83 | 15·7 | 15 | 4·7 |
|  | No | 3 733 733 | 87·0 | 64 776 | 73·1 | 2 082 | 75·7 | 410 | 77·4 | 261 | 81·1 |
|  | Unknown | 226 363 | 5·3 | 6 351 | 7·2 | 182 | 6·6 | 37 | 7·0 | 46 | 14·3 |
| **Vaccine product type (fully vaccinated)** | Comirnaty | 2 494 177 | 67·9 | 5 548 | 71·8 | 158 | 92·4 | 24 | 85·7 | 65 | 97·0 |
|  | Spikevax | 420 588 | 11·4 | 1 038 | 13·4 | 11 | 6·4 | 4 | 14·3 | 2 | 3·0 |
|  | Heterologous mRNA* | 625 060 | 17·0 | 428 | 5·5 | 0 | 0·0 | 0 | 0·0 | 0 | 0·0 |
|  | Vaxzevria<sup>#</sup> | 1 438 | 0·0 | 14 | 0·2 | 0 | 0·0 | 0 | 0·0 | 0 | 0·0 |
|  | Vaxzevria + mRNA vaccine | 132 630 | 3·6 | 702 | 9·1 | 2 | 1·2 | 0 | 0·0 | 0 | 0·0 |

\*combination of vaccine doses of Comirnaty and Spikevax has been administered. <sup>#</sup> Vaxzevria was discontinued in Norway on 11 March 2021, those who received their first dose were offered a second dose of either Comirnaty or Spikevax.

### Overall vaccine effectiveness

The overall effectiveness among fully vaccinated is 72·1 % (71·2 - 73·0) against infection, 92·9 % (91·2 - 94·2) against hospitalisation, adjusted for age, sex, pre-existing medical conditions, living in Oslo/Viken, country of birth, and crowded living conditions (for more model details, see supplementary table S2). Table 2 shows the overall adjusted vaccine effectiveness against SARS-CoV-2 infection, hospitalisation due to COVID-19, admission to ICU and death with COVID-19.

**Table 2.**
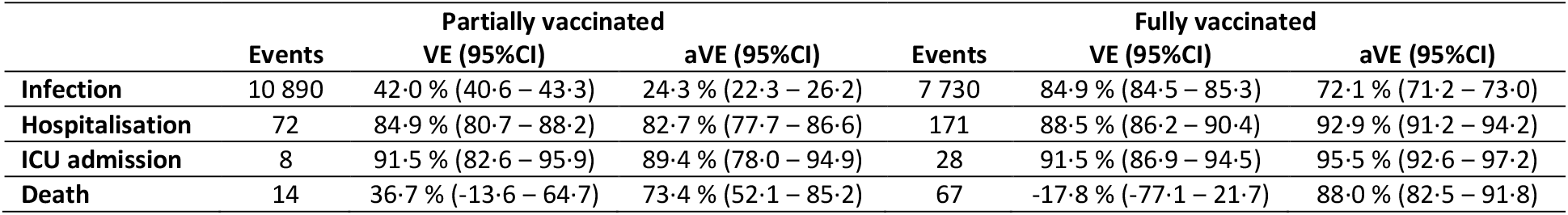
Unadjusted (VE) and adjusted vaccine effectiveness (aVE) against SARS-CoV-2 infection, hospitalisation, ICU admission and death among partially and fully vaccinated individuals in Norway, 1 January 2021 – 27 September 2021. Age, sex, underlying medical conditions, county of residence, country of birth and crowding were adjusted for using stratification.

|  |  | Partially vaccinated |  |  | Fully vaccinated |  |
| --- | --- | --- | --- | --- | --- | --- |
|  | Events | VE (95%CI) | aVE (95%CI) | Events | VE (95%CI) | aVE (95%CI) |
| **Infection** | 10 890 | 42.0 % (40.6 – 43.3) | 24.3 % (22.3 – 26.2) | 7 730 | 84.9 % (84.5 – 85.3) | 72.1 % (71.2 – 73.0) |
| **Hospitalisation** | 72 | 84.9 % (80.7 – 88.2) | 82.7 % (77.7 – 86.6) | 171 | 88.5 % (86.2 – 90.4) | 92.9 % (91.2 – 94.2) |
| **ICU admission** | 8 | 91.5 % (82.6 – 95.9) | 89.4 % (78.0 – 94.9) | 28 | 91.5 % (86.9 – 94.5) | 95.5 % (92.6 – 97.2) |
| **Death** | 14 | 36.7 % (-13.6 – 64.7) | 73.4 % (52.1 – 85.2) | 67 | -17.8 % (-77.1 – 21.7) | 88.0 % (82.5 – 91.8) |

### Vaccine effectiveness by age

Protection against SARS-CoV-2 infection is lower among older age groups, while protection against hospitalization remains reasonably good for all age groups, among both partially and fully vaccinated (Figure 1; supplementary tables S3 and S4).

**Figure 1.**
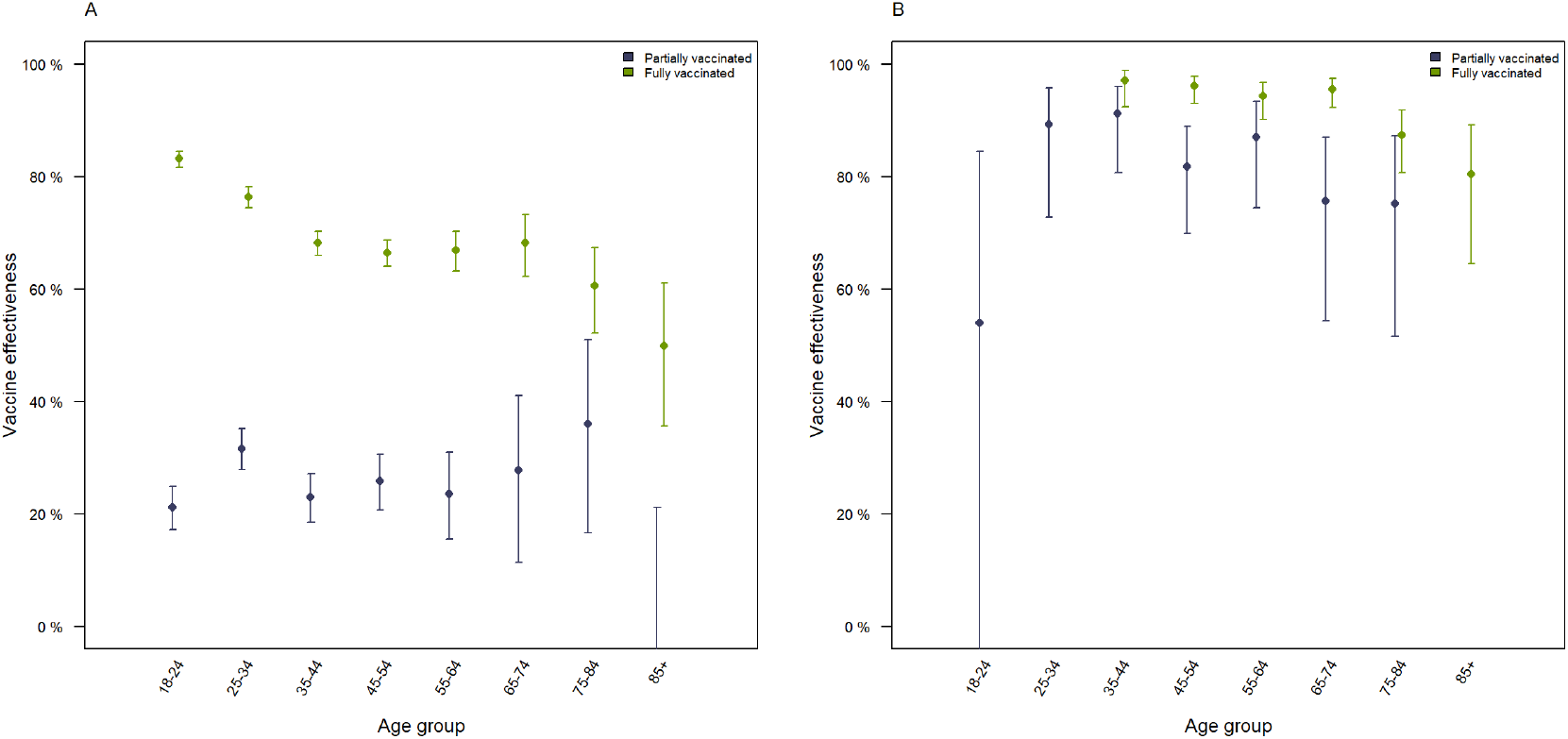
Adjusted vaccine effectiveness against SARS-CoV-2 infection (A) and hospitalisation (B) among partially (blue) and fully (green) vaccinated individuals per age group. Adjusted for sex, pre-existing underlying medical conditions, county of residence, country of birth and crowding.

### Vaccine effectiveness by product type

The vaccine effectiveness against infection among fully vaccinated was highest (84·7 %) for those who received a heterologous mRNA regimen (Figure 2), followed by Spikevax (78·3 %) and Comirnaty (69·7 %), Vaxzevria with mRNA (60·7 %), and two doses of Vaxzevria (43·4 %). Vaccine effectiveness against hospitalisation could only be estimated for Comirnaty and Spikevax and was high: 81·0 % – 84·7 % for partially vaccinated and 91·6 – 96·7 % for fully vaccinated (Figure 2; supplementary tables S5 and S6). There were no hospital admissions among those who received heterologous mRNA vaccination during our study period and too few admissions (<5) among Vaxzevria recipients to reliably infer vaccine effectiveness.

**Figure 2.**
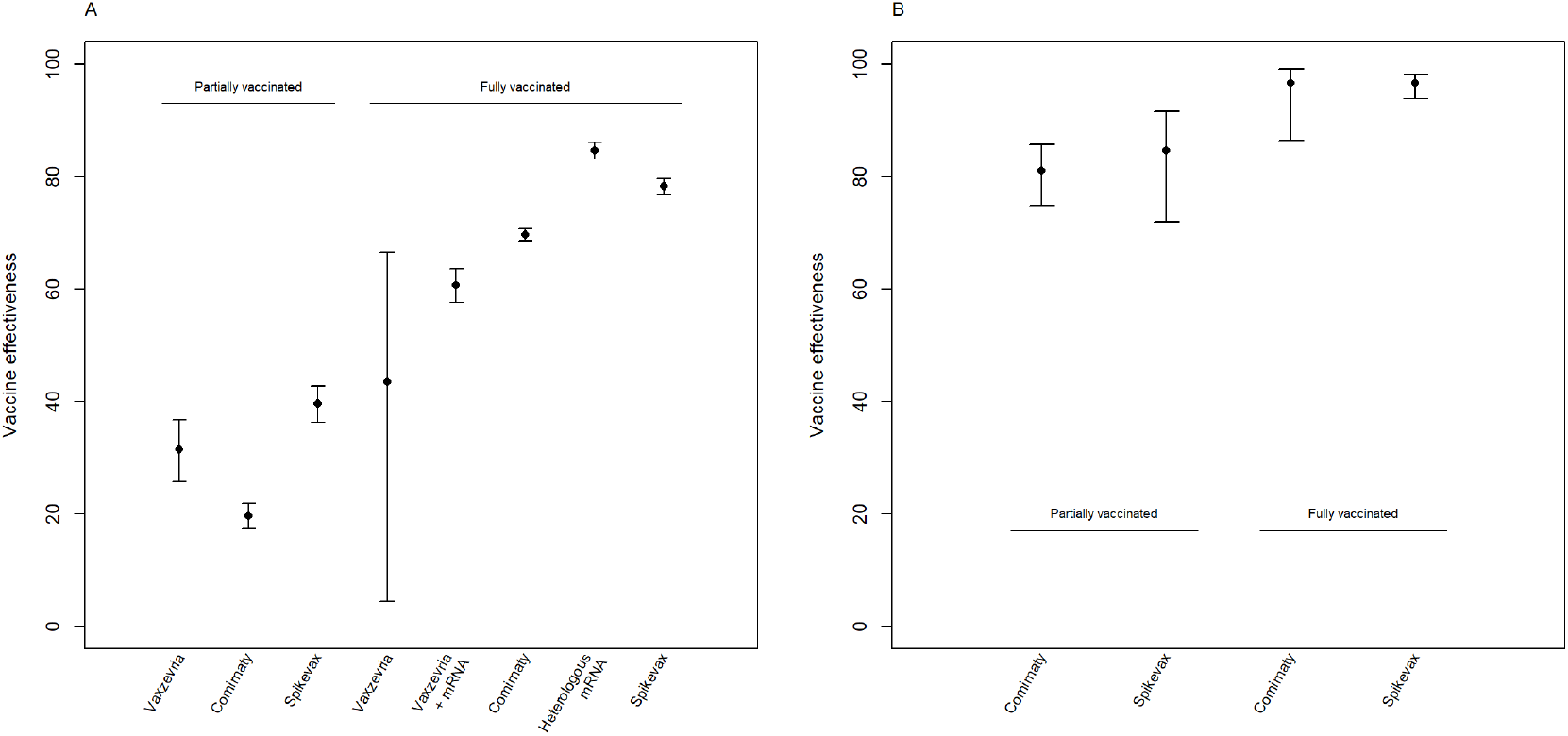
Adjusted vaccine effectiveness against SARS-CoV-2 infection (A) and hospitalisation (B) among partially and fully vaccinated individuals per COVID-19 vaccine product type. Adjusted for age, sex, underlying medical conditions, residence county, country of birth, and crowding.

### Vaccine effectiveness by age and product type

Spikevax showed a higher vaccine effectiveness against infection than Comirnaty in ages <65 years, in those ≥65 years the confidence intervals overlapped. The effectiveness of heterologous mRNA vaccine regimens was as high as, or higher, than either mRNA vaccine separately for the age groups in which it could be estimated. Vaxzevria in combination with mRNA vaccine was less effective against SARS-CoV-2 infection than other vaccine regimens for most age groups (Figure 3, supplementary table S7).

**Figure 3.**
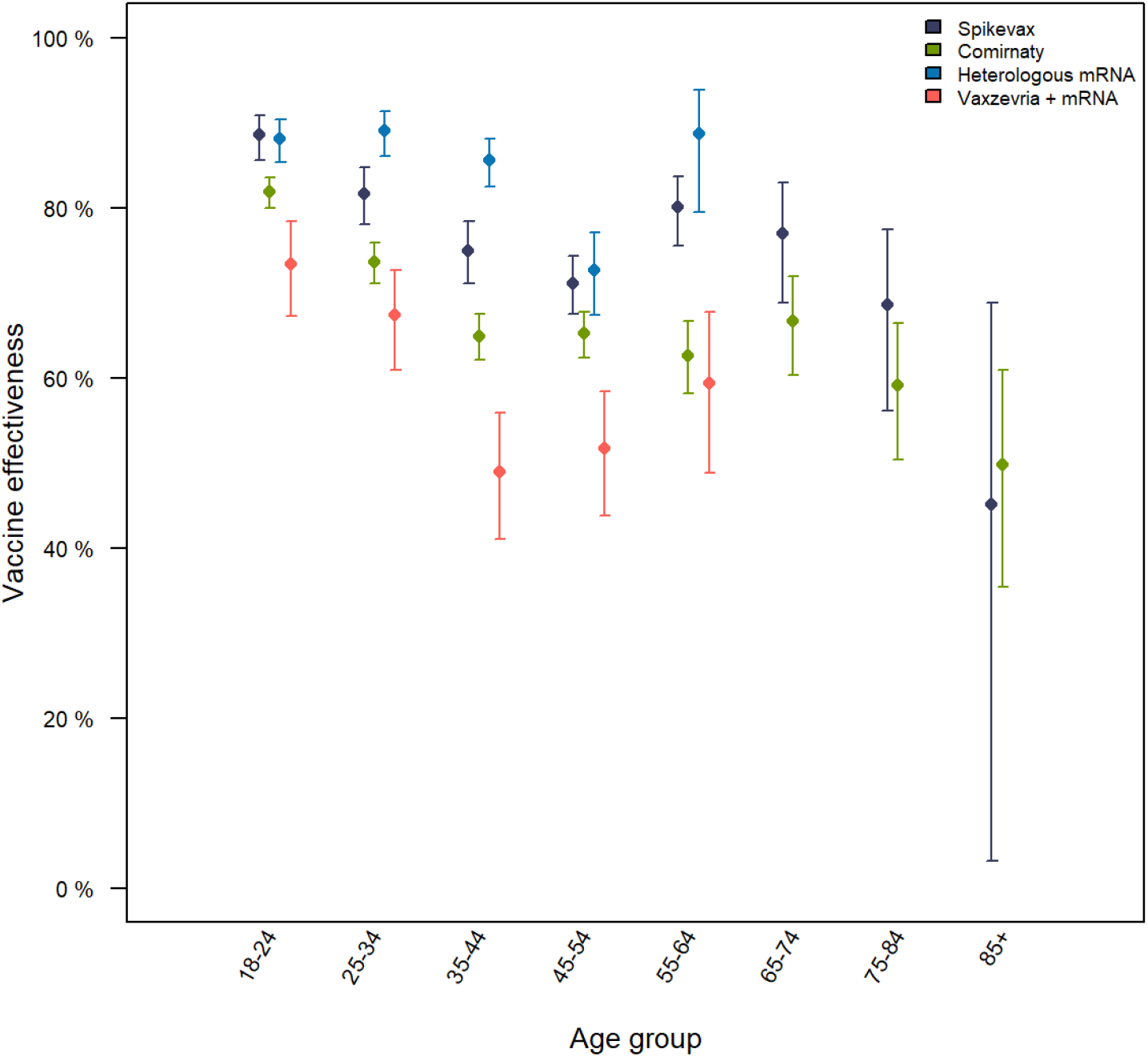
Adjusted vaccine effectiveness (aVE) against SARS-CoV-2 infection by age group and vaccine regimen in Norway, 1^st^ of January – 27^th^ of September 2021. Adjusted for sex, underlying medical conditions, county of residence, country of birth and crowding. For age groups 65 and older, only homologous regimen could be estimated due to zero or low number of cases in heterologous regimen.

## Discussion

Our analysis shows high vaccine effectiveness against SARS-CoV-2 associated hospitalisation, ICU admissions and death associated with COVID-19 among both partially and fully vaccinated individuals, in all age groups and with all vaccine types. However, we found lower protection against SARS-CoV-2 infection among partially vaccinated individuals, as well as a decrease of effectiveness against infection with age among fully vaccinated. In addition, protection against SARS-CoV-2 infection was highest among those who received a homologous Spikevax or heterologous mRNA regimen for the younger age-groups.

To our knowledge, this is the first cohort study to report on the vaccine effectiveness of heterologous mRNA vaccine regimens. As heterologous vaccine regimens were accepted in June, recipients of a mixture of mRNA vaccines are predominantly younger and healthier. However, these results also hold within age groups and are in line with a test-negative case control study from Canada.^21^ In addition, we show that Spikevax shows a higher vaccine effectiveness against infection than Comirnaty, a trend found in all but the oldest age group, as has also been reported by others.^12,22,23^ We found that Vaxzevria in combination with an mRNA vaccine has a lower vaccine effectiveness against infection than the other vaccine regimens. In Norway, Vaxzevria was almost exclusively offered to health care workers, a group with higher risk of exposure and possibly more frequent testing, until 11^th^ of March 2021. As such we might underestimate the effectiveness of Vaxzevria in combination with a mRNA dose due to higher likelihood of finding SARS-CoV-2 infection in this group and by assuming an infection rate among unvaccinated as the general population. Furthermore, only 3·6% of fully vaccinated received this combination regimen, resulting in higher uncertainty of these estimates. Previous studies have shown that both antibody response induced and effectiveness by Vaxzevria in combination with a mRNA vaccine was comparable with homologous mRNA vaccine regimens, but higher than two doses of Vaxzevria.^9,24-26^

Vaccine effectiveness against infection decreased with age, whereas against hospitalisation it remained high with a slight decrease in those over 75 years of age. This difference might be explained by a weaker immune response among older people, the shorter interval between vaccine doses among elderly or waning. In contrast to other studies, we observed lower vaccine effectiveness against death than hospitalisation.^13,22^ The definition used for hospitalisation in this study is possibly more specific than for COVID-19 associated deaths. Comparison of severe disease in terms of hospitalisation and ICU admissions between countries should take into account differences in capacity and structure of health care systems. The hospitals in Norway were not overwhelmed during the study period and therefore the threshold for admission to hospitals might have been lower than in other settings.

During our study period, many aspects of the disease dynamics changed, and changes also occurred simultaneously. This leads to difficulties in correctly identifying the mechanisms and factors driving the system. For example, the introduction of the Delta variant (B.1.617.2) in Norway, coincided with lifting of non-pharmaceutical restrictions, vaccination of younger age groups as well as increasing rates of infection among younger individuals. Due to the prioritisation and roll-out policies of the COVID-19 vaccine program in Norway, various groups have to a certain extent received specific vaccine types at certain times. For example, elderly or those with underlying medical conditions almost exclusively received Comirnaty, whereas Vaxzevria was mainly offered to health care workers. As the heterologous vaccine regimen was accepted in June, recipients of a mixture of mRNA vaccines are predominantly younger and healthier. From the beginning of vaccination, Norway recommended a short interval between doses, extending the interval to 6 – 12 weeks during spring and summer 2021. Studies have demonstrated that longer intervals between doses can result in higher vaccine effectiveness of mRNA vaccines, which again would benefit the younger and healthier groups in our study.^10,27^ In addition, waning could play a role both when looking at the effectiveness by age or vaccine type.^16,28^ Even though we control for these factors in the analyses, residual confounding is to be expected.

Furthermore, testing intensity changed over time, especially with the introduction of self-administered tests. Even though testing capacity is high in Norway, it is likely that not all individuals infected with SARS-CoV-2 have been detected, which could affect the estimates since fraction of unidentified cases might be dependent on age as well as on vaccine status.^29^ Additionally, the estimated vaccine effectiveness can be affected by number and types of contacts.^30^ The vaccine effectiveness could be underestimated if getting vaccinated results in behavioural changes associated with higher risk of exposure. We are not able to model these effects in our analyses, nevertheless the estimated population-level effects can give a reasonable estimate of the individual level vaccine efficacy since the total attack rate during the study period was small.^30^

The ability to link data collected via national registries is a great advantage and allows us to estimate population-wide vaccine effectiveness. However, some limitations of register-based data should be considered when interpreting the results. Data in these registries are not gathered for the purpose of this study, and therefore the focus on level of detail, error checking and precision in the available data is not guided by the current study, as would be the case for independent data gathering. Also, for the combined registries there can be errors for specific individuals, missing individuals, old data and problems when linking individual data from different registries. For example, we adjusted the model for crowded living conditions, which is data collected in 2019 and will be slightly outdated, especially for the younger age groups. And while vaccines administered as part of the Norwegian vaccination program should be in our dataset, it is not unlikely that some individuals (especially those with close ties to other countries) have received vaccines outside Norway and not reported them to SYSVAK.

While changing aspect of disease dynamics leading to bias and limitations in register-based data are important caveats – our cohort study encompassing the whole Norwegian adult population indicates that vaccine effectiveness against severe disease is high among both partially and fully vaccinated individuals. Confounders that could affect our estimates have the highest potential for introducing bias when studying large heterogenous groups. Our estimates remain qualitatively the same for protection against infection and severe disease when splitting by age groups, indicating that the confounding effect of factors that are relatively constant within age-groups introduce little bias in our adjusted models.

Appropriate prioritisation and planning of vaccine campaigns is integral for combating COVID-19 and is only possible with updated knowledge on vaccine effectiveness of realistic vaccination regimens achieved in large populations. Our study of adult Norwegians indicates high protection against severe COVID-19 already after the first vaccine dose. In all regimens where at least one of two doses is an mRNA-vaccine, protection against SARS-CoV-2 infection is good. The results support the use of heterologous schedules, increasing flexibility in vaccination policy.

## Data Availability

Legal restrictions prevent sharing the dataset used in this study. However, researchers are able to request access to linked data from the same registries, as per normal procedure for conducting health research on registry data in Norway. Further information on the preparedness registry, including access to data from each data source, is available at https://www.fhi.no/en/id/infectiousdiseases/coronavirus/emergency-preparedness-register-for-covid-19/

## Authors’ contribution

JS and HM developed the concept and design for the study. JS performed data analyses, TL and GR verified the underlying data and model code. JS, GR, TL, EB, LV, LJ, and HM interpreted the data and drafted the manuscript.

## Data sharing

The datasets analysed during the current study come from the national emergency preparedness registry for COVID-19 (Beredt-C19), housed at the Norwegian Institute of Public Health. The preparedness registry comprises data from a variety of central health registries, national clinical registries, and other national administrative registries. Legal restrictions prevent the researchers from sharing the dataset used in the study. However, external researchers are freely able to request access to linked data from the same registries from outside the structure of Beredt-C19, as per normal procedure for conducting health research on registry data in Norway. Further information on the preparedness registry, including access to data from each data source, is available at https://www.fhi.no/en/id/infectiousdiseases/coronavirus/emergency-preparedness-register-for-covid-19/

## Acknowledgements

The authors greatly appreciate the efforts of all health professionals in vaccinating over 4 million Norwegians and performing over 8 million PCR tests, and in addition managing to report these events to form the data for this analysis. We also acknowledge the efforts of employees at hospitals around Norway in the reporting of timely and complete data to the Norwegian Intensive Care and Pandemic Registry, as well as staff in the registry administration. Additional appreciation is given to all contributors to the development and inner workings of Beredt-C19.

## Supplementary materials

### Data sources and linkages

In this study, we use data from several national registries that were individually linked in the Norwegian preparedness registry (BeredtC19). The data sources and variables used are shown in Table S1

**Table S1.**
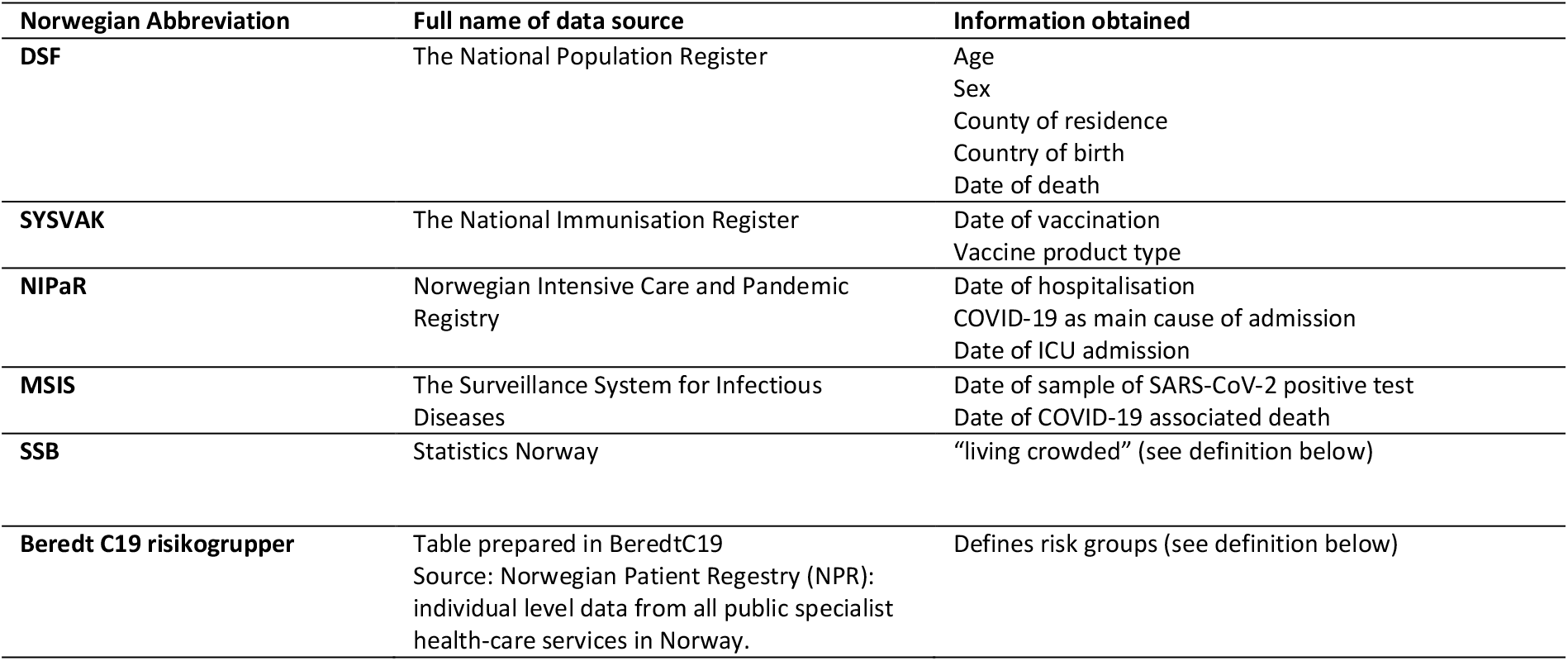
Data sources in the Norwegian preparedness registry (BeredtC19) used in this study and variables retrieved from each source.

#### Crowding^1^

Individuals are consider to live in crowded conditions if the number of rooms is lower than the number of residents or one resident lives in one room, and the number of square metres (P-area) is below 25 sq.m. per person. If the number of rooms or the P-area is not specified, a household will be regarded as living in cramped conditions if one of these criteria is met.

#### Risk groups

Some underlying medical conditions increase the risk of severe COVID-19 outcomes, regardless of age. In Norway, these individuals have been prioritised in the vaccination campaigns. The underlying comorbidities that have been defined as increasing the risk of severe COVID-19 are divided into two groups:

High risk: people with diseases/conditions that carry a high risk of severe COVID-19:

- Organ transplant
- Immunodeficiency
- Hematological cancer in the last five years
- Other active cancers
- Neurological or neuromuscular diseases that cause impaired cough or lung function (e.g., ALS and cerebral palsy)
- Chronic kidney disease, or significant renal impairment.

Medium risk: people with diseases/conditions that entail a moderate risk of severe COVID-19:

- Chronic liver disease or significant hepatic impairment
- Diseases requiring immunosuppressive therapy
- Diabetes
- Chronic lung disease including cystic fibrosis and severe asthma which have required the use of high dose inhaled or oral steroids within the past year
- Obesity with a body mass index (BMI) of ≥35 kg/m2
- Dementia
- Chronic heart and vascular disease (except for high blood pressure) and stroke

### The COVID-19 epidemic in Norway Jan-Sep 2021

Figure S1 shows the number of infections and hospitalisation from January 1^st^ to September 27^th^, 2021. Note that these are the data included in our study, and numbers can deviate slightly from other reported numbers due to data cleaning and exclusion as described in the main manuscript. Figure S2 shows the changing proportion of the different vaccine status categories from January 1^st^ to September 27^th^, 2021.

**Figure S1.**
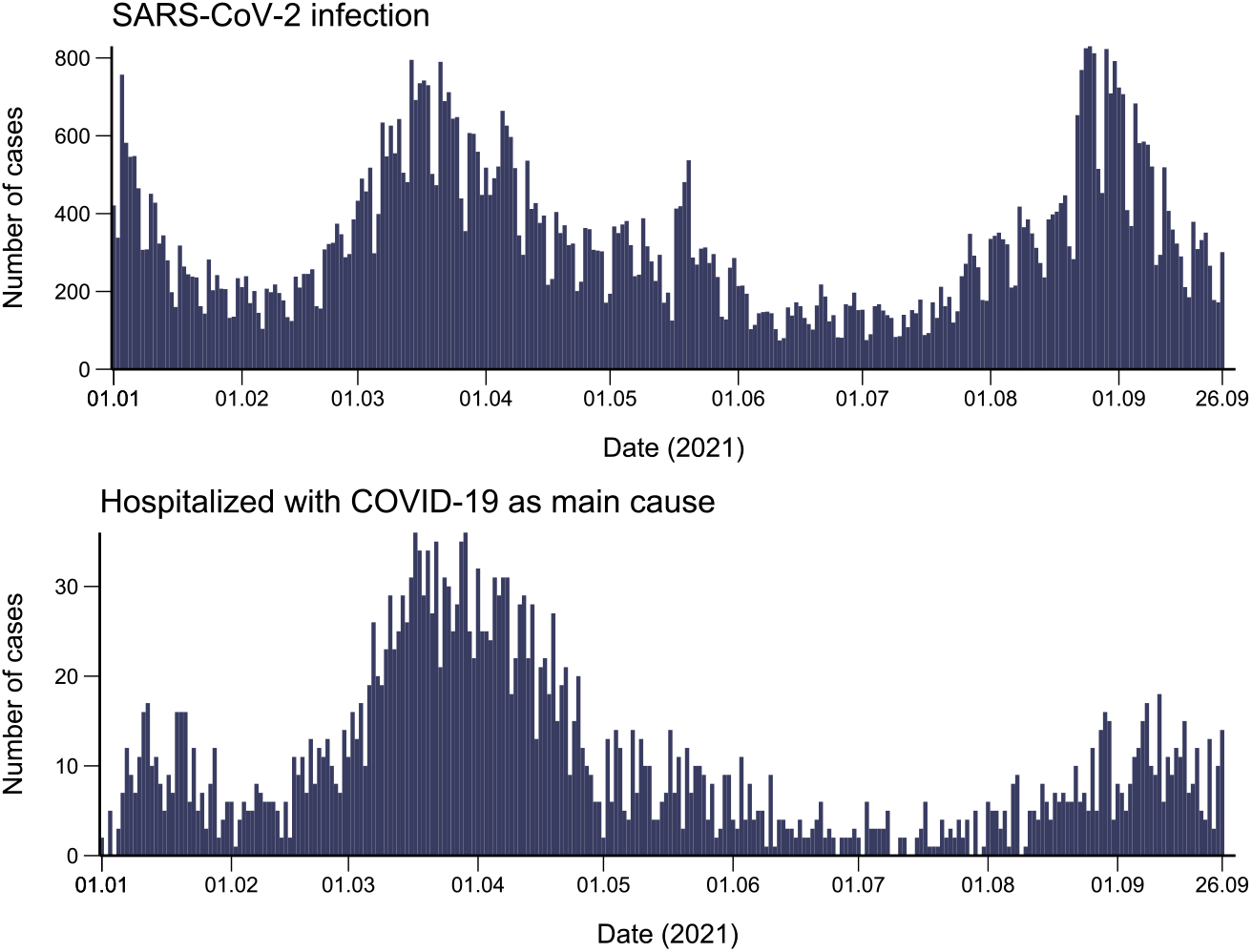
Reported cases of SARS-CoV-2 infection, and number of hospitalisations in Norway, 1 January -27 September 2021.

**Figure S2.**
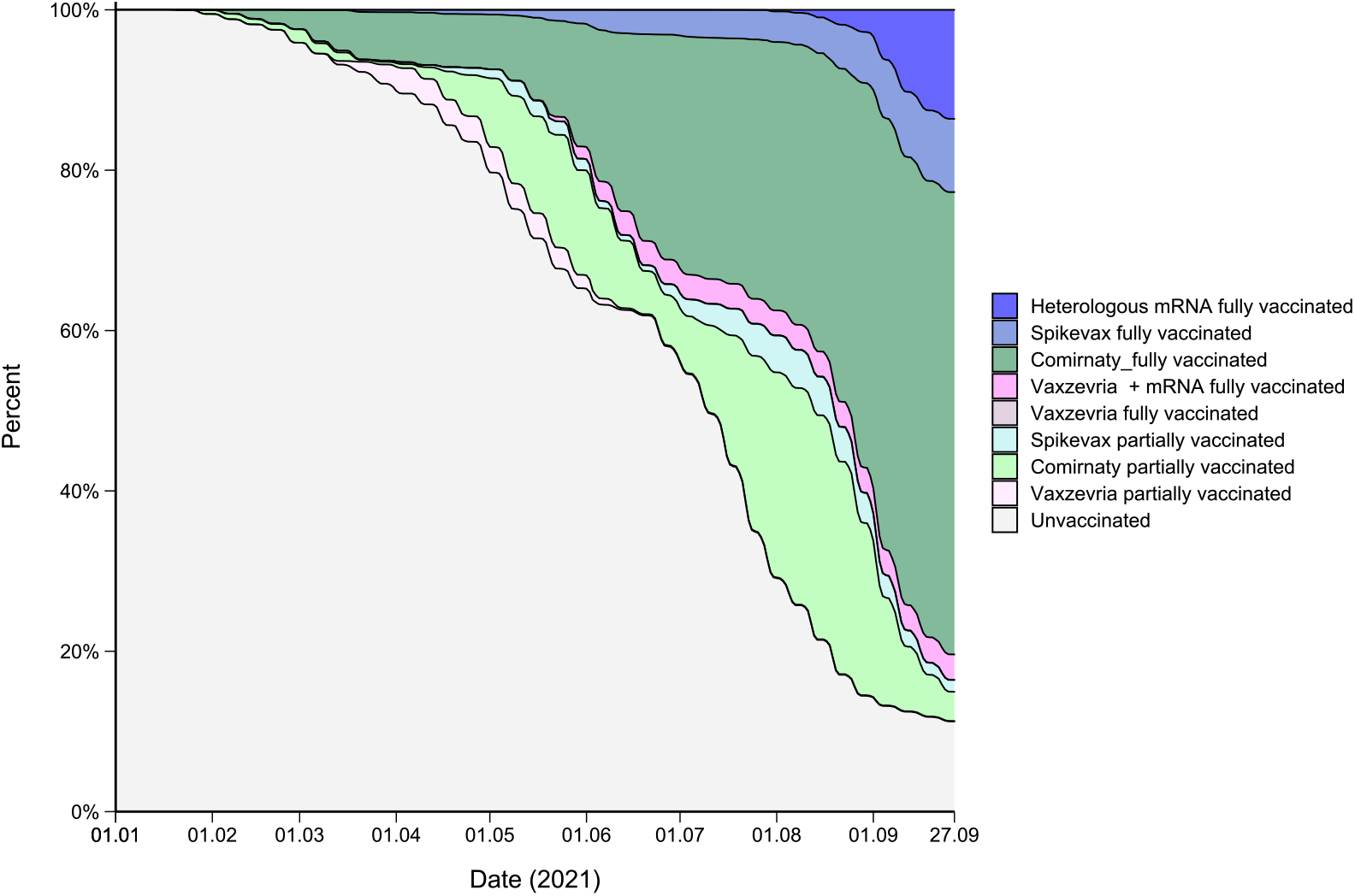
Proportion of COVID-19 vaccine product type in Norway, 1st January -27th September 2021.

### Modelling overall vaccine effectiveness

Using Cox proportional hazard models, we estimated the COVID-19 vaccine effectiveness against SARS-CoV-2 infection, hospitalisation, ICU admission, and death associated with COVID-19. We ran three sets of model structures:

1. Unadjusted models only using time-varying vaccine status as covariate. This yields crude vaccine effectiveness estimates.
2. Adjusted models with vaccine status as well as age, sex, crowding and risk group as fixed covariates assuming proportional impact on hazard rates, and using county of residence and country of birth as stratification variables. These were performed to test for impact of covariates.
3. Adjusted models with all covariates, except vaccine status, implemented as stratification variables. This is the most general way to adjust for the covariates allowing each combination of covariate groups to have their own baseline hazard rate, while still assuming that vaccination leads to a proportional change in risk of infection.

The reported adjusted vaccine effectiveness estimates in the manuscript are from the third model structure allowing impacts of all covariates except vaccine status to possibly be non-proportional. Results from model structure 2 is presented in table S2. Note that the youngest age-group was used as reference in all models, and as there are 0 deaths in this age-group the proportional hazards for age-groups in model for the mortality outcome are approaching infinity and are nonsensical. However, since the model uses partial likelihoods, the parameter estimates for the other covariates are still meaningful. We also checked this by running a separate Cox model with age group 65 – 74 as reference, giving identical estimates for the other parameters. Adjustment assuming proportionality in covariates age, sex, risk-group, and crowding lead to vaccine effectiveness (Table S2) which are very similar to adjustments assuming different baseline hazard-rates using the *strata* functionality in *survival*, (Table 2, main manuscript).

**Table S2.**
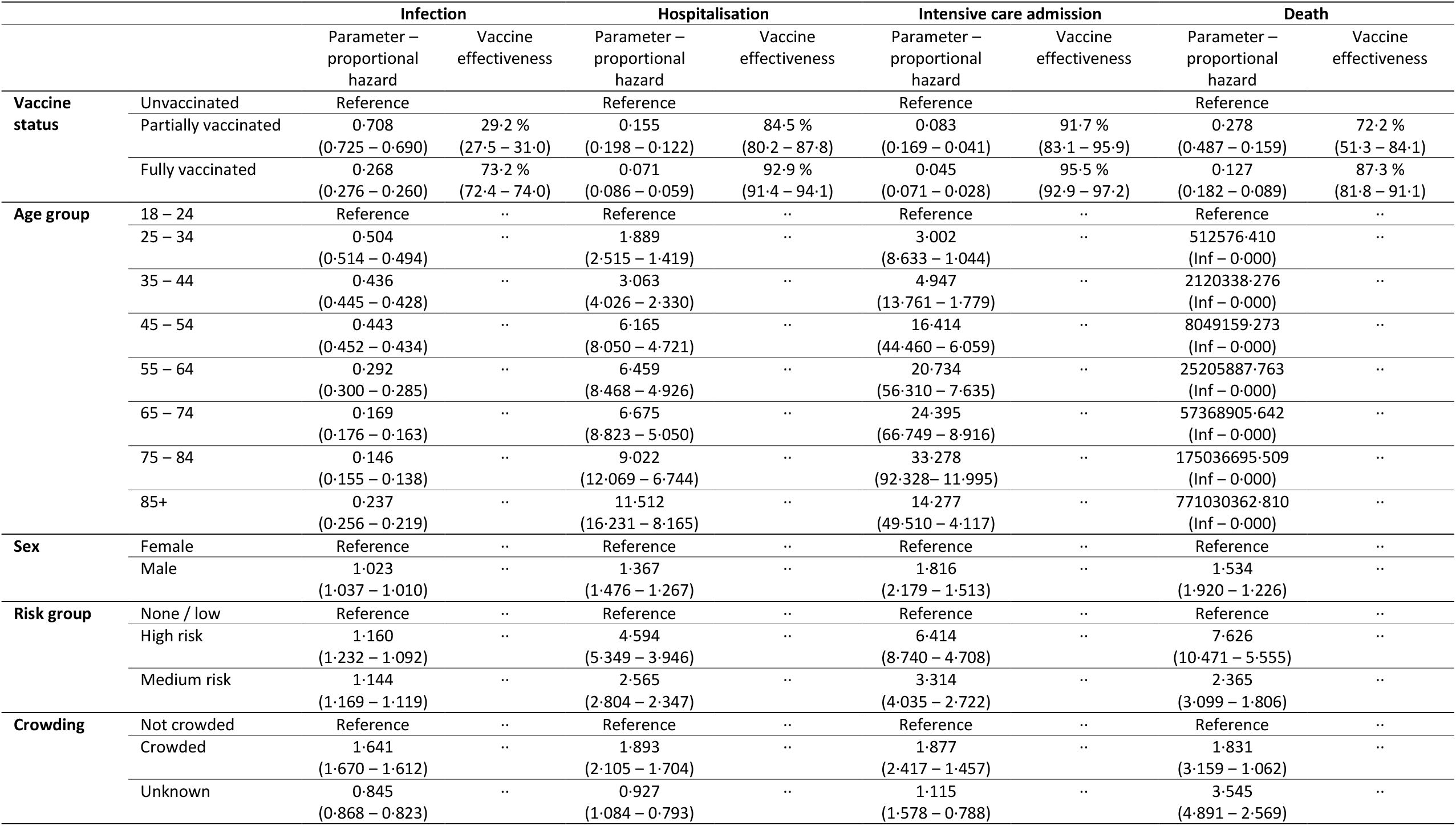
Proportional effects in hazard rates of SARS-CoV-2 infection, hospitalisation, ICU admission and death associated with COVID-19 individuals in Norway, 1 January -27 September 2021. Vaccine effectiveness (100 * (1 – *β*)) is also given for vaccine status.

### Vaccine effectiveness stratified by age and product type

The estimates used for figure 1 (stratified by age) in the manuscript are shown in Table S3 (infection) and S4 (hospitalisation).

**Table S3.**
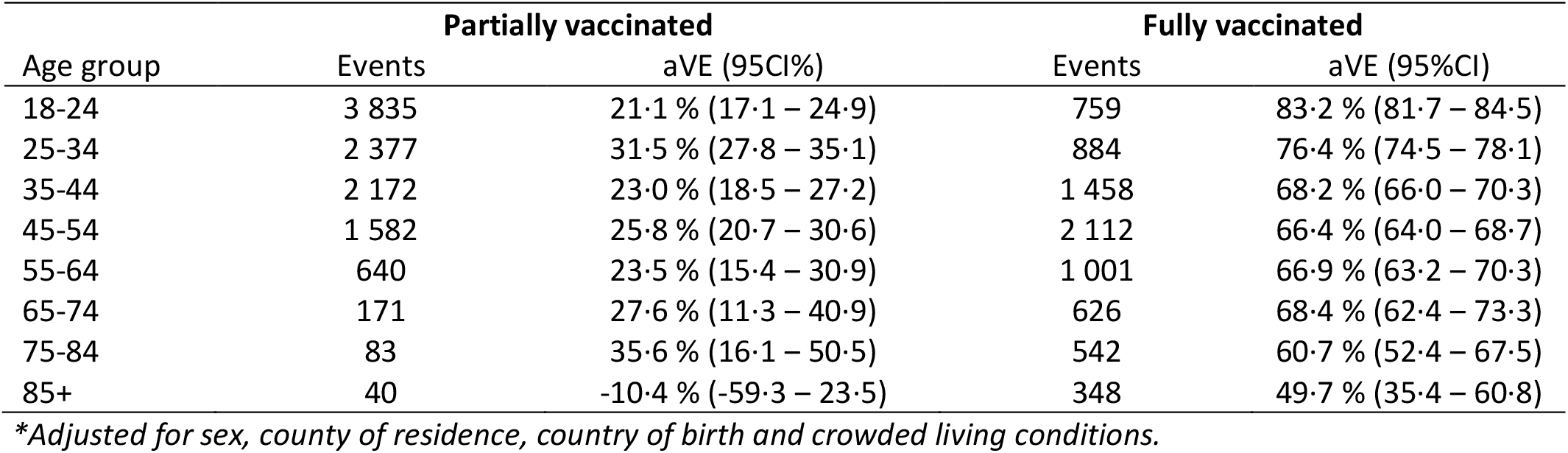
Adjusted vaccine effectiveness (aVE)* against SARS-CoV-2 infection for partially and fully vaccinated individuals by age in Norway, 1 January -27 September 2021.

**Table S4.**
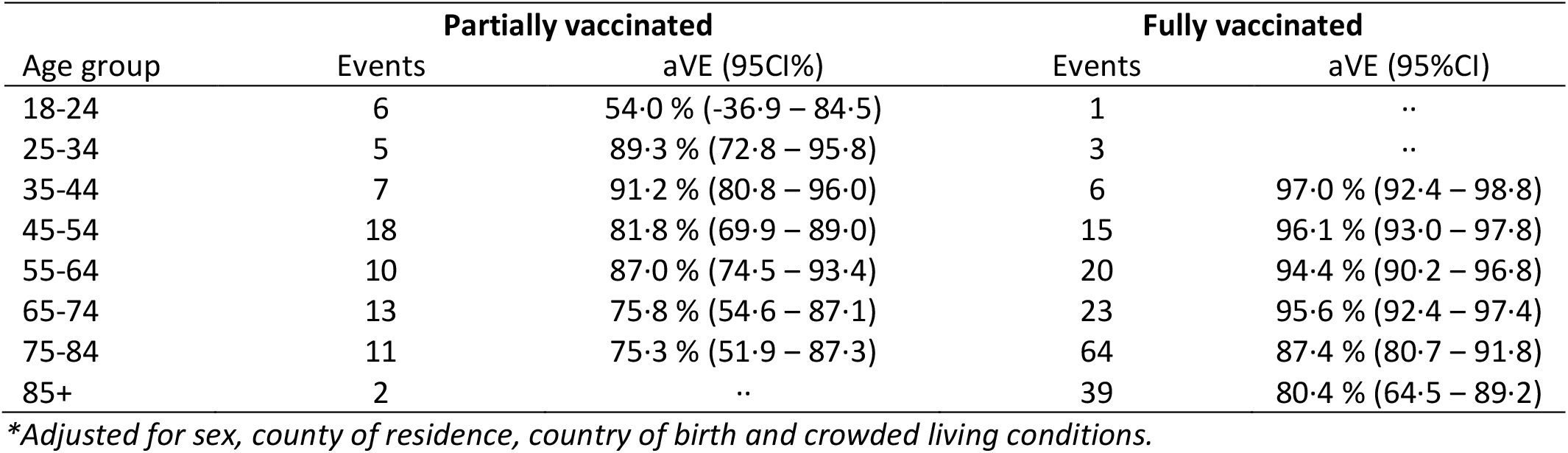
Adjusted vaccine effectiveness (aVE)* against hospitalisation due to COVID-19 for partially and fully vaccinated individuals by age in Norway, 1 January -27 September 2021.

The estimates used for figure 2 (stratified by product type) in the manuscript are shown in Table S5 (infection) and S6 (hospitalisation).

**Table S5.**
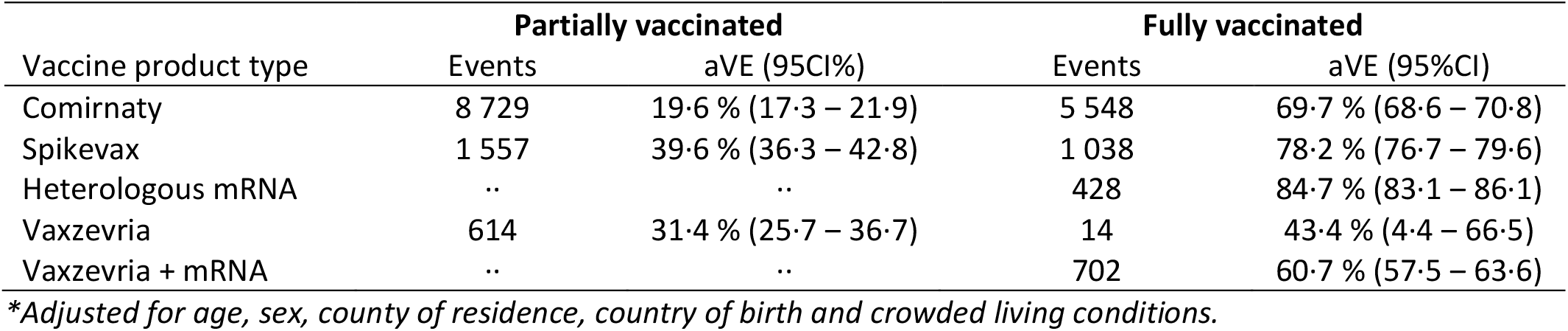
Adjusted vaccine effectiveness (aVE)* against SARS-CoV-2 infection for partially and fully vaccinated individuals by vaccine product type in Norway, 1 January -27 September 2021.

**Table S6.**
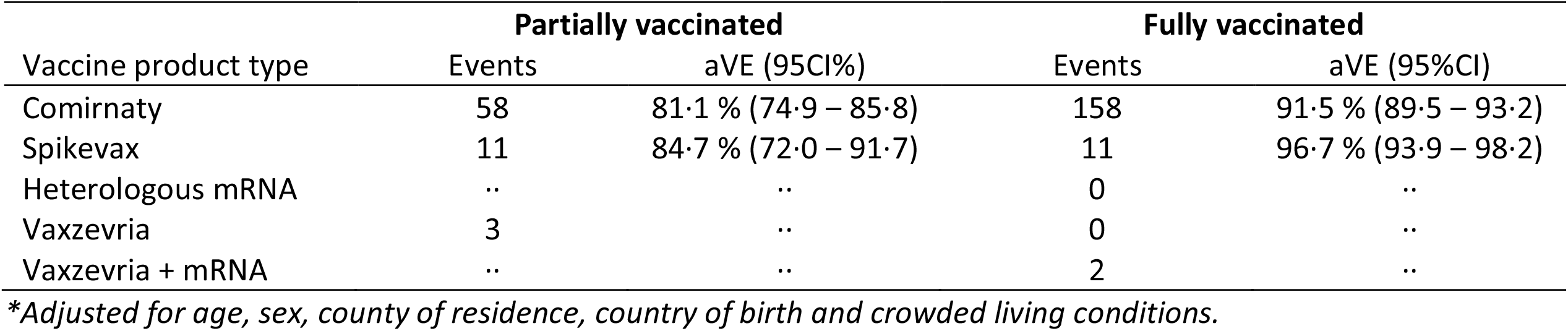
Adjusted vaccine effectiveness (aVE)* against hospitalisation due to COVID-19 for partially and fully vaccinated individuals by vaccine product type in Norway, 1 January -27 September 2021.

Table S7 shows the adjusted vaccine effectiveness for partially and fully vaccinated by age and vaccine product type.

**Table S7.**
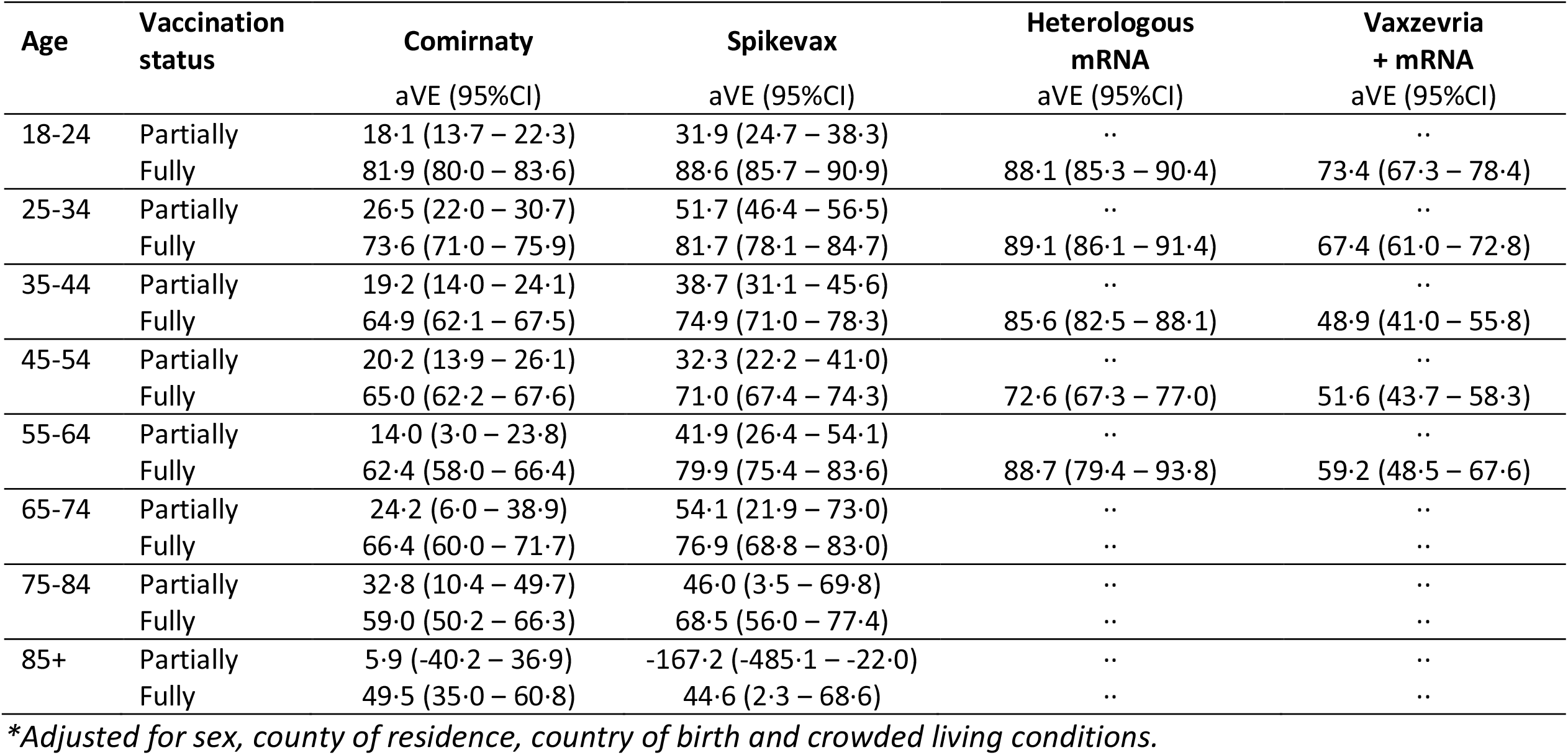
Adjusted vaccine effectiveness(aVE)* against SARS-CoV-2 infection partially and fully vaccinated individuals by age and vaccine product type in Norway, 1 January -27 September 2021.

In addition to age and product type, we also stratified by risk group to estimate vaccine effectiveness against infection (figure S3).

**Figure S3.**
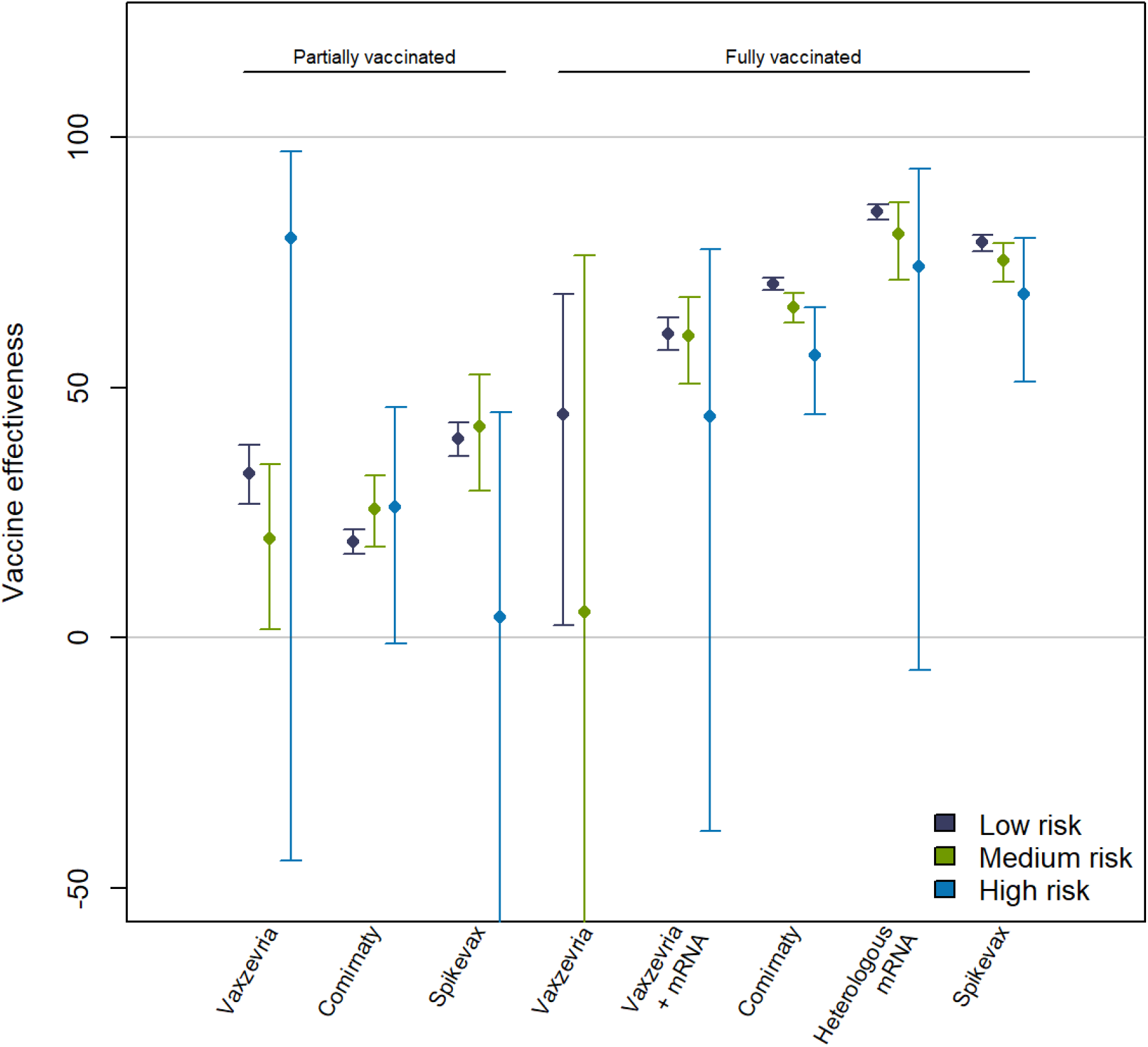
Estimated COVID-19 vaccine effectiveness against SARS-CoV-2 infection by risk group and product type in Norway, 1 January -27 September 2021. *Risk groups are determined based on underlying medical conditions with increased risk of severe disease, classified as high, medium, and low risk*.

