## Supplementary material for "Age and product dependent vaccine effectiveness against SARS-CoV-2 infection and hospitalisation among adults in Norway: a national cohort study, January – September 2021"

### Supplementary materials
#### Data sources and linkages
In this study, we use data from several national registries that were individually linked in the Norwegian preparedness registry (BeredtC19). The data sources and variables used are shown in Table S1

**Table S1. Data sources in the Norwegian preparedness registry (BeredtC19) used in this study and variables retrieved from each source**

| Norwegian Abbreviation | Full name of data source | Information obtained |
|---|---|---|
| **DSF** | The National Population Register | Age<br>Sex<br>County of residence<br>Country of birth<br>Date of death |
| **SYSVAK** | The National Immunisation Register | Date of vaccination<br>Vaccine product type |
| **NIPaR** | Norwegian Intensive Care and Pandemic Registry | Date of hospitalisation<br>COVID-19 as main cause of admission<br>Date of ICU admission |
| **MSIS** | The Surveillance System for Infectious Diseases | Date of sample of SARS-CoV-2 positive test<br>Date of COVID-19 associated death |
| **SSB** | Statistics Norway | "living crowded" (see definition below) |
| **Beredt C19 risikogrupper** | Table prepared in BeredtC19<br>Source: Norwegian Patient Regestry (NPR): individual level data from all public specialist health-care services in Norway. | Defines risk groups (see definition below) |

Crowding[1]: Individuals are consider to live in crowded conditions if the number of rooms is lower than the number of residents or one resident lives in one room, and the number of square metres (P-area) is below 25 sq.m. per person. If the number of rooms or the P-area is not specified, a household will be regarded as living in cramped conditions if one of these criteria is met.

Risk groups: Some underlying medical conditions increase the risk of severe COVID-19 outcomes, regardless of age. In Norway, these individuals have been prioritised in the vaccination campaigns. The underlying comorbidities that have been defined as increasing the risk of severe COVID-19 are divided into two groups:

High risk: people with diseases/conditions that carry a high risk of severe COVID-19:
- Organ transplant
- Immunodeficiency
- Hematological cancer in the last five years
- Other active cancers
- Neurological or neuromuscular diseases that cause impaired cough or lung function (e.g., ALS and cerebral palsy)
- Chronic kidney disease, or significant renal impairment.

Medium risk: people with diseases/conditions that entail a moderate risk of severe COVID-19:
- Chronic liver disease or significant hepatic impairment
- Diseases requiring immunosuppressive therapy
- Diabetes
- Chronic lung disease including cystic fibrosis and severe asthma which have required the use of high dose inhaled or oral steroids within the past year
- Obesity with a body mass index (BMI) of ≥35 kg/m2
- Dementia
- Chronic heart and vascular disease (except for high blood pressure) and stroke

**The COVID-19 epidemic in Norway Jan-Sep 2021**

Figure S1 shows the number of infections and hospitalisation from January 1st to September 27th, 2021. Note that these are the data included in our study, and numbers can deviate slightly from other reported numbers due to data cleaning and exclusion as described in the main manuscript. Figure S2 shows the changing proportion of the different vaccine status categories from January 1st to September 27th, 2021.

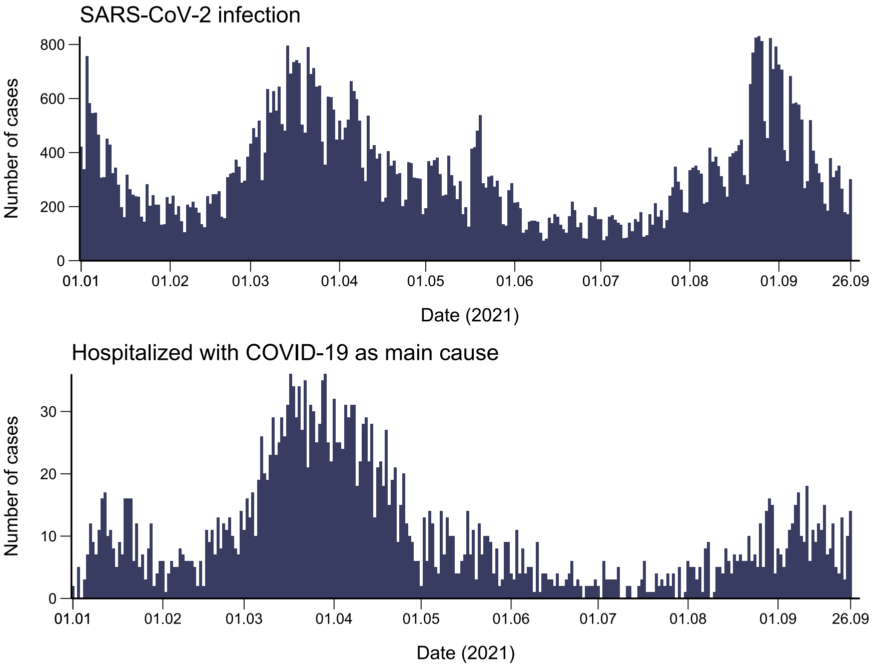


**Figure S1. Reported cases of SARS-CoV-2 infection, and number of hospitalisations in Norway, 1 January -27 September 2021**

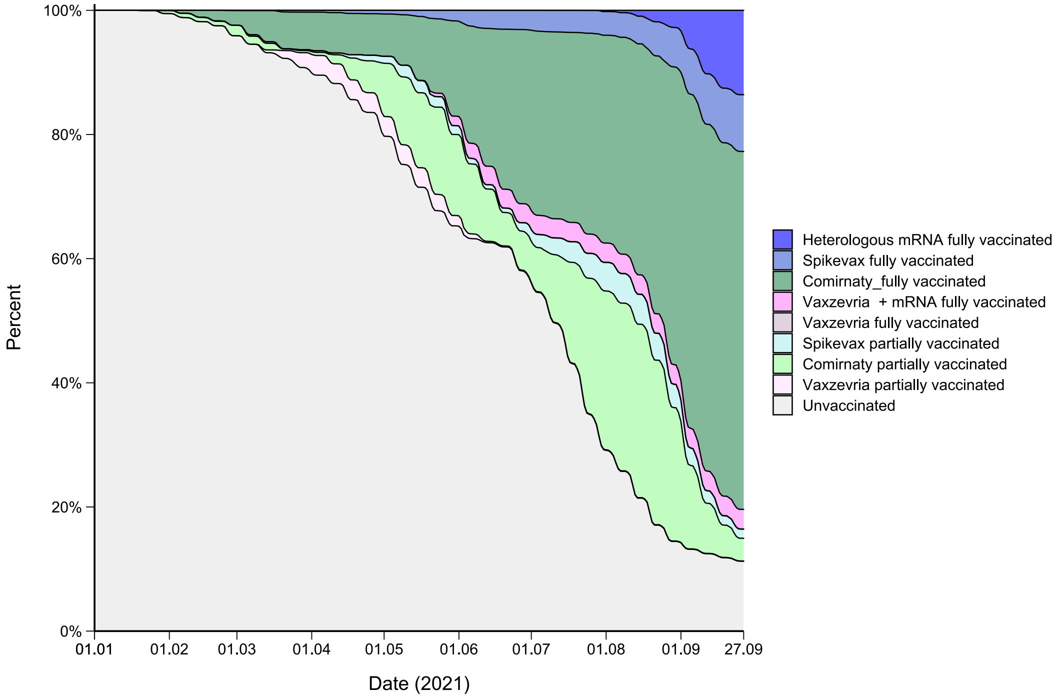


**Figure S2. Proportion of COVID-19 vaccine product type in Norway, 1st January -27th September 2021.**

**Table S2. Proportional effects in hazard rates of SARS-CoV-2 infection, hospitalisation, ICU admission and death associated with COVID-19 individuals in Norway, 1 January -27 September 2021. Vaccine effectiveness (100 * (1 – *β*)) is also given for vaccine status.**

| | | **Infection** | | **Hospitalisation** | | **Intensive care admission** | | **Death** | |
|---|---|---|---|---|---|---|---|---|---|
| | | Parameter – proportional hazard | Vaccine effectiveness | Parameter – proportional hazard | Vaccine effectiveness | Parameter – proportional hazard | Vaccine effectiveness | Parameter – proportional hazard | Vaccine effectiveness |
| **Vaccine status** | Unvaccinated | Reference | | Reference | | Reference | | Reference | |
| | Partially vaccinated | 0·708 (0·725 – 0·690) | 29·2 % (27·5 – 31·0) | 0·155 (0·198 – 0·122) | 84·5 % (80·2 – 87·8) | 0·083 (0·169 – 0·041) | 91·7 % (83·1 – 95·9) | 0·278 (0·487 – 0·159) | 72·2 % (51·3 – 84·1) |
| | Fully vaccinated | 0·268 (0·276 – 0·260) | 73·2 % (72·4 – 74·0) | 0·071 (0·086 – 0·059) | 92·9 % (91·4 – 94·1) | 0·045 (0·071 – 0·028) | 95·5 % (92·9 – 97·2) | 0·127 (0·182 – 0·089) | 87·3 % (81·8 – 91·1) |
| **Age group** | 18 – 24 | Reference | ·· | Reference | ·· | Reference | ·· | Reference | ·· |
| | 25 – 34 | 0·504 (0·514 – 0·494) | ·· | 1·889 (2·515 – 1·419) | ·· | 3·002 (8·633 – 1·044) | ·· | 512576·410 (Inf – 0·000) | ·· |
| | 35 – 44 | 0·436 (0·445 – 0·428) | ·· | 3·063 (4·026 – 2·330) | ·· | 4·947 (13·761 – 1·779) | ·· | 2120338·276 (Inf – 0·000) | ·· |
| | 45 – 54 | 0·443 (0·452 – 0·434) | ·· | 6·165 (8·050 – 4·721) | ·· | 16·414 (44·460 – 6·059) | ·· | 8049159·273 (Inf – 0·000) | ·· |
| | 55 – 64 | 0·292 (0·300 – 0·285) | ·· | 6·459 (8·468 – 4·926) | ·· | 20·734 (56·310 – 7·635) | ·· | 25205887·763 (Inf – 0·000) | ·· |
| | 65 – 74 | 0·169 (0·176 – 0·163) | ·· | 6·675 (8·823 – 5·050) | ·· | 24·395 (66·749 – 8·916) | ·· | 57368905·642 (Inf – 0·000) | ·· |
| | 75 – 84 | 0·146 (0·155 – 0·138) | ·· | 9·022 (12·069 – 6·744) | ·· | 33·278 (92·328– 11·995) | ·· | 175036695·509 (Inf – 0·000) | ·· |
| | 85+ | 0·237 (0·256 – 0·219) | ·· | 11·512 (16·231 – 8·165) | ·· | 14·277 (49·510 – 4·117) | ·· | 771030362·810 (Inf – 0·000) | ·· |
| **Sex** | Female | Reference | ·· | Reference | ·· | Reference | ·· | Reference | ·· |
| | Male | 1·023 (1·037 – 1·010) | ·· | 1·367 (1·476 – 1·267) | ·· | 1·816 (2·179 – 1·513) | ·· | 1·534 (1·920 – 1·226) | ·· |
| **Risk group** | None / low | Reference | ·· | Reference | ·· | Reference | ·· | Reference | ·· |
| | High risk | 1·160 (1·232 – 1·092) | ·· | 4·594 (5·349 – 3·946) | ·· | 6·414 (8·740 – 4·708) | ·· | 7·626 (10·471 – 5·555) | ·· |
| | Medium risk | 1·144 (1·169 – 1·119) | ·· | 2·565 (2·804 – 2·347) | ·· | 3·314 (4·035 – 2·722) | ·· | 2·365 (3·099 – 1·806) | ·· |
| **Crowding** | Not crowded | Reference | ·· | Reference | ·· | Reference | ·· | Reference | ·· |
| | Crowded | 1·641 (1·670 – 1·612) | ·· | 1·893 (2·105 – 1·704) | ·· | 1·877 (2·417 – 1·457) | ·· | 1·831 (3·159 – 1·062) | ·· |
| | Unknown | 0·845 (0·868 – 0·823) | ·· | 0·927 (1·084 – 0·793) | ·· | 1·115 (1·578 – 0·788) | ·· | 3·545 (4·891 – 2·569) | ·· |

**Vaccine effectiveness stratified by age and product type**

The estimates used for figure 1 (stratified by age) in the manuscript are shown in Table S3 (infection) and S4 (hospitalisation).

**Table S3. Adjusted vaccine effectiveness (aVE)* against SARS-CoV-2 infection for partially and fully vaccinated individuals by age in Norway, 1 January -27 September 2021**

| | Partially vaccinated | | Fully vaccinated | |
|---|---|---|---|---|
| Age group | Events | aVE (95CI%) | Events | aVE (95%CI) |
| 18-24 | 3 835 | 21·1 % (17·1 – 24·9) | 759 | 83·2 % (81·7 – 84·5) |
| 25-34 | 2 377 | 31·5 % (27·8 – 35·1) | 884 | 76·4 % (74·5 – 78·1) |
| 35-44 | 2 172 | 23·0 % (18·5 – 27·2) | 1 458 | 68·2 % (66·0 – 70·3) |
| 45-54 | 1 582 | 25·8 % (20·7 – 30·6) | 2 112 | 66·4 % (64·0 – 68·7) |
| 55-64 | 640 | 23·5 % (15·4 – 30·9) | 1 001 | 66·9 % (63·2 – 70·3) |
| 65-74 | 171 | 27·6 % (11·3 – 40·9) | 626 | 68·4 % (62·4 – 73·3) |
| 75-84 | 83 | 35·6 % (16·1 – 50·5) | 542 | 60·7 % (52·4 – 67·5) |
| 85+ | 40 | -10·4 % (-59·3 – 23·5) | 348 | 49·7 % (35·4 – 60·8) |

**Adjusted for sex, county of residence, country of birth and crowded living conditions.*

**Table S4. Adjusted vaccine effectiveness (aVE)* against hospitalisation due to COVID-19 for partially and fully vaccinated individuals by age in Norway, 1 January -27 September 2021**

| | Partially vaccinated | | Fully vaccinated | |
|---|---|---|---|---|
| Age group | Events | aVE (95CI%) | Events | aVE (95%CI) |
| 18-24 | 6 | 54·0 % (-36·9 – 84·5) | 1 | ·· |
| 25-34 | 5 | 89·3 % (72·8 – 95·8) | 3 | ·· |
| 35-44 | 7 | 91·2 % (80·8 – 96·0) | 6 | 97·0 % (92·4 – 98·8) |
| 45-54 | 18 | 81·8 % (69·9 – 89·0) | 15 | 96·1 % (93·0 – 97·8) |
| 55-64 | 10 | 87·0 % (74·5 – 93·4) | 20 | 94·4 % (90·2 – 96·8) |
| 65-74 | 13 | 75·8 % (54·6 – 87·1) | 23 | 95·6 % (92·4 – 97·4) |
| 75-84 | 11 | 75·3 % (51·9 – 87·3) | 64 | 87·4 % (80·7 – 91·8) |
| 85+ | 2 | ·· | 39 | 80·4 % (64·5 – 89·2) |

**Adjusted for sex, county of residence, country of birth and crowded living conditions.*

The estimates used for figure 2 (stratified by product type) in the manuscript are shown in Table S5 (infection) and S6 (hospitalisation).

**Table S5. Adjusted vaccine effectiveness (aVE)* against SARS-CoV-2 infection for partially and fully vaccinated individuals by vaccine product type in Norway, 1 January -27 September 2021**

| | Partially vaccinated | | Fully vaccinated | |
|---|---|---|---|---|
| Vaccine product type | Events | aVE (95CI%) | Events | aVE (95%CI) |
| Comirnaty | 8 729 | 19·6 % (17·3 – 21·9) | 5 548 | 69·7 % (68·6 – 70·8) |
| Spikevax | 1 557 | 39·6 % (36·3 – 42·8) | 1 038 | 78·2 % (76·7 – 79·6) |
| Heterologous mRNA | ·· | ·· | 428 | 84·7 % (83·1 – 86·1) |
| Vaxzevria | 614 | 31·4 % (25·7 – 36·7) | 14 | 43·4 % (4·4 – 66·5) |
| Vaxzevria + mRNA | ·· | ·· | 702 | 60·7 % (57·5 – 63·6) |

**Adjusted for age, sex, county of residence, country of birth and crowded living conditions.*

**Table S6. Adjusted vaccine effectiveness (aVE)* against hospitalisation due to COVID-19 for partially and fully vaccinated individuals by vaccine product type in Norway, 1 January -27 September 2021**

| | **Partially vaccinated** | | **Fully vaccinated** | |
|---|---|---|---|---|
| Vaccine product type | Events | aVE (95CI%) | Events | aVE (95%CI) |
| Comirnaty | 58 | 81·1 % (74·9 – 85·8) | 158 | 91·5 % (89·5 – 93·2) |
| Spikevax | 11 | 84·7 % (72·0 – 91·7) | 11 | 96·7 % (93·9 – 98·2) |
| Heterologous mRNA | ·· | ·· | 0 | ·· |
| Vaxzevria | 3 | ·· | 0 | ·· |
| Vaxzevria + mRNA | ·· | ·· | 2 | ·· |

**Adjusted for age, sex, county of residence, country of birth and crowded living conditions.*

Table S7 shows the adjusted vaccine effectiveness for partially and fully vaccinated by age and vaccine product type.

**Table S7. Adjusted vaccine effectiveness(aVE)* against SARS-CoV-2 infection partially and fully vaccinated individuals by age and vaccine product type in Norway, 1 January -27 September 2021**

| **Age** | **Vaccination status** | **Comirnaty** | **Spikevax** | **Heterologous mRNA** | **Vaxzevria + mRNA** |
|---|---|---|---|---|---|
| | | aVE (95%CI) | aVE (95%CI) | aVE (95%CI) | aVE (95%CI) |
| 18-24 | Partially | 18·1 (13·7 – 22·3) | 31·9 (24·7 – 38·3) | ·· | ·· |
| | Fully | 81·9 (80·0 – 83·6) | 88·6 (85·7 – 90·9) | 88·1 (85·3 – 90·4) | 73·4 (67·3 – 78·4) |
| 25-34 | Partially | 26·5 (22·0 – 30·7) | 51·7 (46·4 – 56·5) | ·· | ·· |
| | Fully | 73·6 (71·0 – 75·9) | 81·7 (78·1 – 84·7) | 89·1 (86·1 – 91·4) | 67·4 (61·0 – 72·8) |
| 35-44 | Partially | 19·2 (14·0 – 24·1) | 38·7 (31·1 – 45·6) | ·· | ·· |
| | Fully | 64·9 (62·1 – 67·5) | 74·9 (71·0 – 78·3) | 85·6 (82·5 – 88·1) | 48·9 (41·0 – 55·8) |
| 45-54 | Partially | 20·2 (13·9 – 26·1) | 32·3 (22·2 – 41·0) | ·· | ·· |
| | Fully | 65·0 (62·2 – 67·6) | 71·0 (67·4 – 74·3) | 72·6 (67·3 – 77·0) | 51·6 (43·7 – 58·3) |
| 55-64 | Partially | 14·0 (3·0 – 23·8) | 41·9 (26·4 – 54·1) | ·· | ·· |
| | Fully | 62·4 (58·0 – 66·4) | 79·9 (75·4 – 83·6) | 88·7 (79·4 – 93·8) | 59·2 (48·5 – 67·6) |
| 65-74 | Partially | 24·2 (6·0 – 38·9) | 54·1 (21·9 – 73·0) | ·· | ·· |
| | Fully | 66·4 (60·0 – 71·7) | 76·9 (68·8 – 83·0) | ·· | ·· |
| 75-84 | Partially | 32·8 (10·4 – 49·7) | 46·0 (3·5 – 69·8) | ·· | ·· |
| | Fully | 59·0 (50·2 – 66·3) | 68·5 (56·0 – 77·4) | ·· | ·· |
| 85+ | Partially | 5·9 (-40·2 – 36·9) | -167·2 (-485·1 – -22·0) | ·· | ·· |
| | Fully | 49·5 (35·0 – 60·8) | 44·6 (2·3 – 68·6) | ·· | ·· |

**Adjusted for sex, county of residence, country of birth and crowded living conditions.*

In addition to age and product type, we also stratified by risk group to estimate vaccine effectiveness against infection (figure S3).

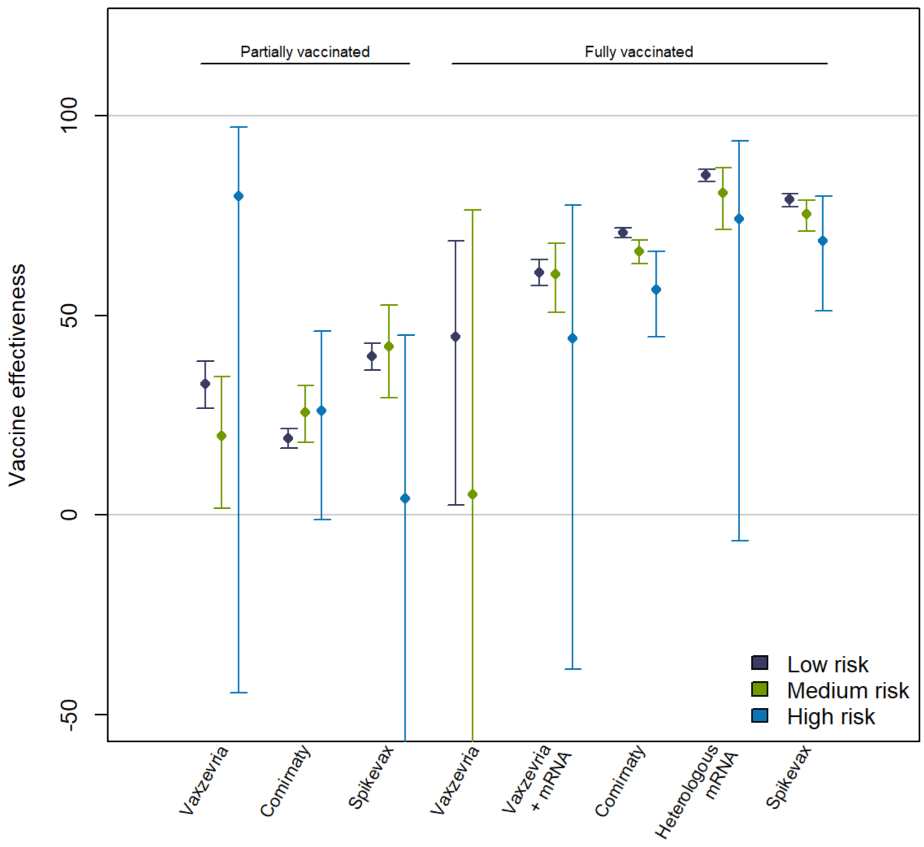


**Figure S3. Estimated COVID-19 vaccine effectiveness against SARS-CoV-2 infection by risk group and product type in Norway, 1 January -27 September 2021.** *Risk groups are determined based on underlying medical conditions with increased risk of severe disease, classified as high, medium, and low risk.*

1. Statistics Norway. Concept variable: Crowded dwelling. 2020. https://www.ssb.no/a/metadata/conceptvariable/vardok/3462/en (accessed 18.10 2021).
